# Reducing opioid use for chronic pain with mobile health interventions: a systematic scoping review

**DOI:** 10.1101/2025.08.07.25333246

**Authors:** Michael R Magee, Ali Gholamrezaei, Amy G McNeilage, Alison Sim, Paul Glare, Claire E Ashton-James

## Abstract

This systematic scoping review explores the extent and characteristics of research on mobile health (mHealth) interventions for reducing opioid use in chronic pain management. A comprehensive search was run on eight major bibliographic databases (e.g., PubMed) as well as grey literature (e.g., clinical trials registries). Each record was screened by two independent reviewers. Studies were included if they investigated an mHealth intervention for adults with chronic pain and reported opioid use as an outcome (primary or secondary). Data of the study characteristics and results (if available) were extracted and analysed descriptively. Out of 3097 records, 25 (11 published studies, 14 protocols) were included. The studies were from five countries and conducted between 2009-2023. mHealth Interventions included mobile applications (20), text messaging (4), and interactive voice response (1). In about half of the studies (13), taking opioids was an eligibility criterion. In four studies, interventions were specifically designed to support tapering. Studies with published results concluded that the mHealth interventions are acceptable to participants (9) and generally feasible (6). This systematic scoping review shows that there is a growing interest in research investigating mHealth interventions to support people with chronic pain tapering opioids. Current research mostly investigated mobile applications designed for chronic pain management with opioids use as a secondary outcome. mHealth interventions specifically designed to support opioids tapering are emerging. Available results show that mHealth interventions are acceptable, feasible, and potentially efficacious for patients with chronic pain tapering opioids.

## Introduction

Chronic pain, pain lasting longer than 3 months^1^, is the greatest cause of years lost to disability globally^2,3^ and a huge financial drain on society^4,5^. Opioid medications are frequently prescribed for chronic pain but pose significant safety concerns, particularly with long-term use, including tolerance, dependence, cognitive impairment, and potentially fatal respiratory depression^6-8^. Patients currently taking prescribed opioid medications for whom the risks outweigh the benefits are advised to reduce or discontinue their use of opioid medications with their clinician, a process known as tapering^8-13^. However, tapering is challenging for many patients, who can experience negative impacts on pain, mood, and uncomfortable withdrawal symptoms^6,7,13,14^.

Patients who have successfully tapered report that a combination of pain education, opioid tapering support, and social support (from clinicians and personal networks) may facilitate tapering success^15-19^. A systematic review and meta-analysis investigating the efficacy of interventions to reduce long-term opioid therapy for chronic pain concluded that the provision of multidisciplinary support such as psychology and physiotherapy pain self-management programs, is probably effective in reducing opioid doses^20^. However, multidisciplinary support is costly, and most pain treatment services cannot meet demand^21^. This inaccessibility adversely impacts those living with chronic pain and may perpetuate long-term opioid medication use when it is ill advised^13,22^.

Digital health may provide a scalable, inexpensive solution to the lack of accessibility for opioid tapering support^23^. Digital platforms such as mobile phones and other portable technology provide patients with accessible support for many chronic health issues. Digital health supports may improve health equity and address the inequitable distribution of chronic pain treatment options across geographic and socioeconomic gradients^21,24,25^. A range of innovative digital interventions are either in use or in development to support chronic pain management^26-29^. For example, a systematic review of digital health interventions concluded there is good support for the use of internet-delivered interventions for chronic pain^30^. Flexible, at home digital solutions may be a particularly good fit within a chronic pain population, due to the levels of physical disability and associated challenges that some people living with chronic pain may experience^25^. Further, the stigma that some people with chronic pain report around opioid use may be mitigated through the anonymity of digital health supports^31,32^.

Evidence regarding mobile health (mHealth) interventions to support patients with chronic pain tapering opioids is emerging. However, based on a preliminary search in March 2023 of MEDLINE, the Cochrane Database of Systematic Reviews, JBI Evidence Synthesis, and PROSPERO databases, there is currently no synthesised evidence (published or protocol) in this area. Therefore, this scoping review was conducted to address the primary research question ‘what is the scope of available evidence for mHealth in providing support to individuals with chronic pain during opioid tapering?’. Several review sub-questions were developed to align data extraction and reporting with the areas of interest for the review (Table 1).

**Table 1:** Research Questions.

| Primary Research Question |
| --- |
| What is the scope of available evidence for mHealth to support opioid tapering for individuals with chronic pain? |
| Secondary Research Questions |
| In what geographical and clinical contexts (research settings) have the mHealth interventions included in the review been delivered? |
| What are the characteristics of the study sample populations? |
| What are the characteristics of the mHealth interventions in the reviewed studies? |
| What is the degree to which consumer involvement or co-design is included in the reviewed studies? |
| What is the scope of evidence on acceptability and feasibility of interventions in the reviewed studies? |
| What is the degree to which theoretical frameworks and clinical interventions underpin the design and or content of mHealth interventions? |

## Methods

### Protocol

This review was informed by the Joanne Briggs Institute (JBI) Reviewer’s manual for the conduct and reporting of scoping reviews^34^ as well as the Preferred Reporting Items for Systematic Reviews and Meta-Analysis Extension for Scoping Reviews (PRISMA-ScR)^33^. A scoping review protocol was registered on Open Science Framework in February 2023^35^. A PRISMA-ScR checklist and summary of changes to the protocol after registration are included in the supplemental digital content.

### Population, Concept, Context (PCC)

To define the scope of this review, a population, concept, context (PCC) framework was developed. The population considers studies of adults living with chronic pain. The two concepts are mHealth and opioids. The concept of mHealth is mobile phone-based digital health technologies for the purpose of delivering healthcare support or an intervention to patients^36,37^. Specifically, mHealth supports are asynchronous to clinician care (i.e. not digital telehealth services or didactic phone calls). The concept of opioids refers to any prescribed opioid medication for the management of chronic pain. Regarding context, studies were limited to research conducted since 1990. This timepoint should capture the inclusion of when mHealth technologies were increasingly accessible to the public. The inclusion and exclusion criteria are described in Table 2.

**Table 2:** Inclusion and Exclusion Criteria.

| **Inclusion Criteria** |
| --- |
| Studies must investigate a mobile health intervention for adults (18+) who have chronic non-cancer pain. |
| The intervention must be asynchronous to clinician care (i.e. not telehealth). |
| Participants must be prescribed opioids for chronic pain (i.e., not acute, cancer, or surgical pain). |
| Participants must receive an mHealth intervention to either support opioid tapering (or reducing reliance on opioid medications), or opioid dose is reported as an outcome within a more general chronic pain mHealth intervention. |
| Studies with both qualitative and quantitative methods will be included. Studies must be written in English. |
| **Exclusion Criteria** |
| Participants do not report chronic pain (e.g., the study is for opioid use disorder only or participants do not experience chronic pain). |
| Studies in which opioid use or dose is measured as a baseline variable only. |
| Animal studies. |
| Studies which use SMS or App's only for appointment management. |
| Studies that use mobile phones to measure study outcomes only (e.g. SMS is used to administer an outcome measure). |
| Interventions that are delivered to healthcare providers. |
| Studies published before the year 1990. |
| Duplicate research articles reporting data related to the same study (data can be extracted from both for completeness). |
| Protocols which later had data published in another included study. |

### Information Sources

The scoping review considers both experimental and quasi-experimental study designs including randomised/non-randomised controlled trials, before and after studies, and interrupted time-series studies. In addition, analytical observational studies including prospective and retrospective cohort studies, case-control studies, and analytical cross-sectional studies are included. Descriptive observational study designs including case series, individual case reports, and descriptive cross-sectional studies and qualitative studies that met other inclusion criteria were included. Systematic and other reviews, as well as text and opinion papers, were included if they reported original data. Grey literature included conference presentations, theses, and clinical trial protocols.

### Search Strategy

A database search strategy was developed by the researchers. Keywords linked to the PCC of chronic pain, opioids, and mHealth, were developed based on related review articles^20,30,38^. The search was run on PubMed (including MEDLINE), Scopus, Embase, CINAHL, APA PsychInfo, Global Health, JBI EBP Database, and the Cochrane Library. For grey literature, searches were run on ProQuest Dissertations and Theses, EBSCO Open Dissertations, The Conference Proceedings Citation Index (via Web of Science), and the World Health Organisation (WHO) Clinical Trials Registry. The search strategy, including all keywords, medical subject headings, and index terms were adapted for each included database or information source. One researcher carried out all database searches in April 2023. The search strategy is included in the supplemental digital content. A second search (in the same databases) was conducted in October 2024 to find any reported results of the clinical trials protocols included in the review. Authors of the clinical trials protocols were contacted if no published result was found.

### Study Selection

Results were uploaded to Covidence^39^ to manage screening. The title and abstract of each source (first pass) and then the full text (second pass) were screened by two independent reviewers. Disagreements were resolved through discussion or with a third reviewer. If there were multiple results for the same study, such as a trial protocol then the published findings, or the publication of the same data in more than one article/report, the earlier publications were excluded. Despite excluding earlier versions, any additional data that could contribute to the completeness of the review was also extracted.

### Pilot Search

A pilot search was conducted to test the search, screening, and extraction procedure. A sample of 100 articles was selected from an initial search. The two reviewers had a high rate of agreement (above 75% agreement) considered acceptable in the protocol^34^. Eligible texts from the pilot search were extracted using the charting table from the protocol and changes were made to refine the extraction table template.

### Data Extraction and Data Analysis

A descriptive analytic approach was utilised for data analysis^40^. Data relevant to the review questions and sub questions were extracted from each article and reported descriptively. One reviewer extracted the data from each study which was then verified by another member of the research team. In line with our aims and established guidelines for scoping reviews, we did not complete a quality assessment of the results of each study in this review^41 34^.

## Results

### What is the scope of available evidence for mHealth to support opioid tapering for individuals with chronic pain?

The searches identified 3097 results from which 25 articles were included. See Figure 1 for the search flow including major reasons for exclusion. Eleven articles reported the results of original investigations^42-52^ (hereafter referred to as research studies) and 14 reported protocols. Of the research studies, five were randomised clinical trials (RCTs)^44,45,49,50,52^, four were non-randomised published studies ^42,43,46,51^, and two published abstracts from conference presentations (one RCT, one single-arm study)^47,48^. One published article^53^ excluded at full text review (due to no opioid outcomes being reported) used the same sample and data as a conference abstract that was included^47^. The conference article reported opioid outcomes and was more recent, so was retained, though some data was drawn from the more comprehensive article^47,53^. The remaining 14 articles were trial protocols^54 55 56 57 58 59 51 60 61 62 63 64 65 66,67^. Five of the protocols were published in peer-reviewed journals^55 46 58 60 59^ and the others were drawn from clinical trial registries^54,56,61-67^. If protocols were published both in articles and trial registries, the article was cited^59 68^. Of the 14 protocols, 11 were protocols for RCTs ^55 56 57 59 51 60 61 63 64 65 66,67^ while three ^54,58,62^ described single-arm open label trials. The extracted research studies and protocols were published between 2009 and 2023. There were a wide range of reported study durations, from two-weeks to up to a year. Sample sizes in the research studies ranged from 18 to 2245 participants. Of the 14 protocols, target sample sizes ranged from 40 to 350 participants and two did not report target sample sizes^54,62^. The results discussed below are presented in Table 3.

**Figure 1:**
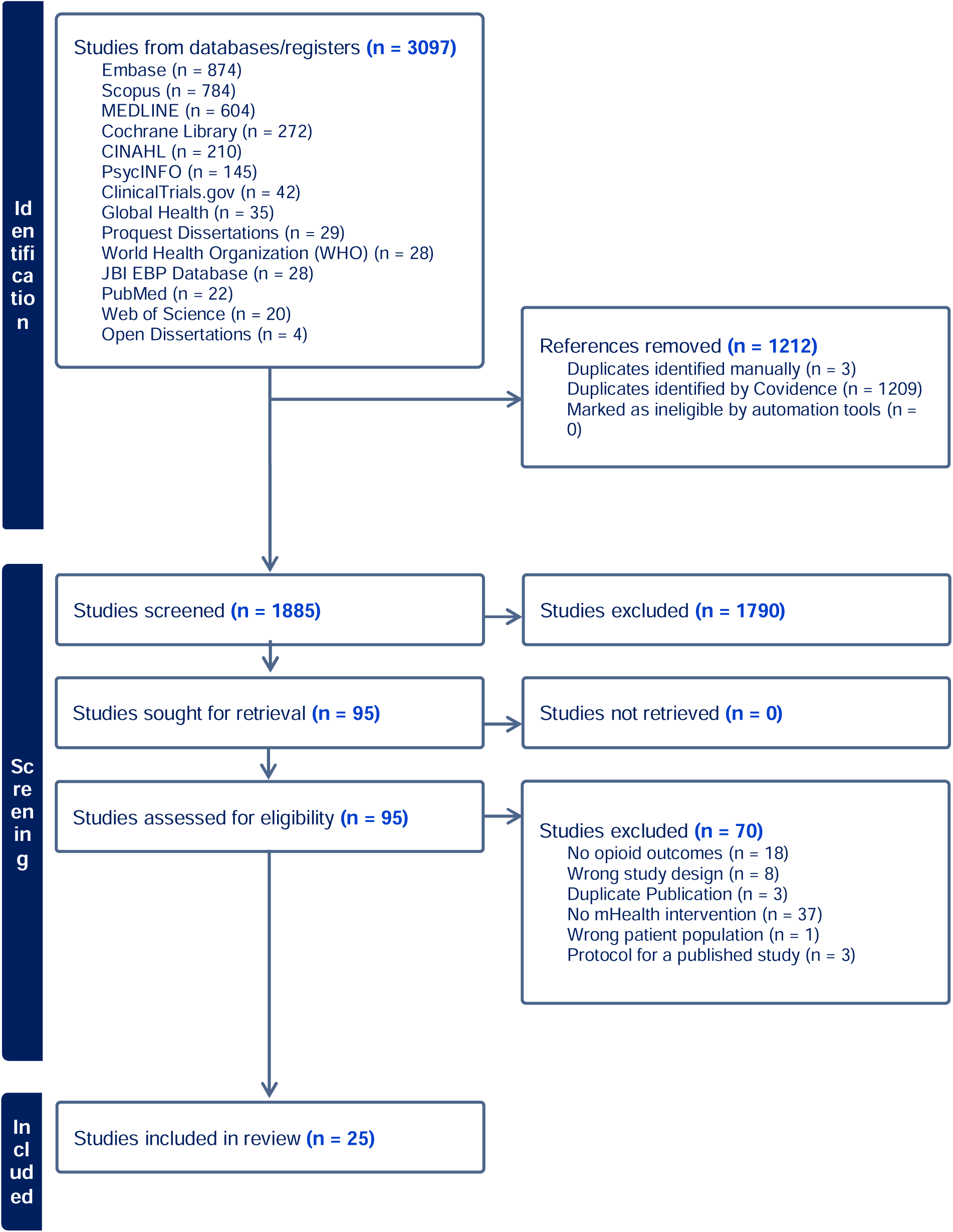
PRISMA Study Flow Chart.

**Table 3.**
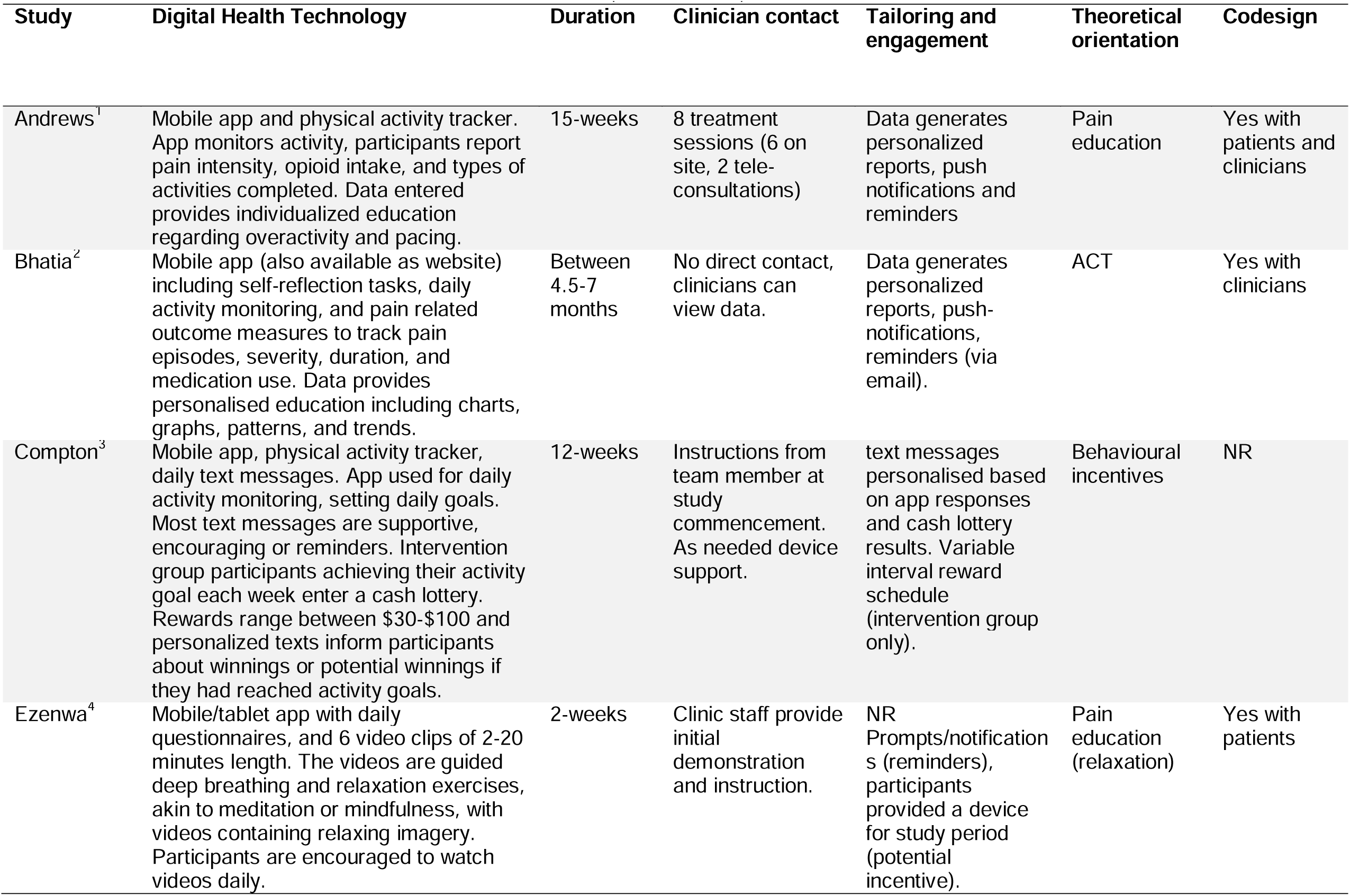
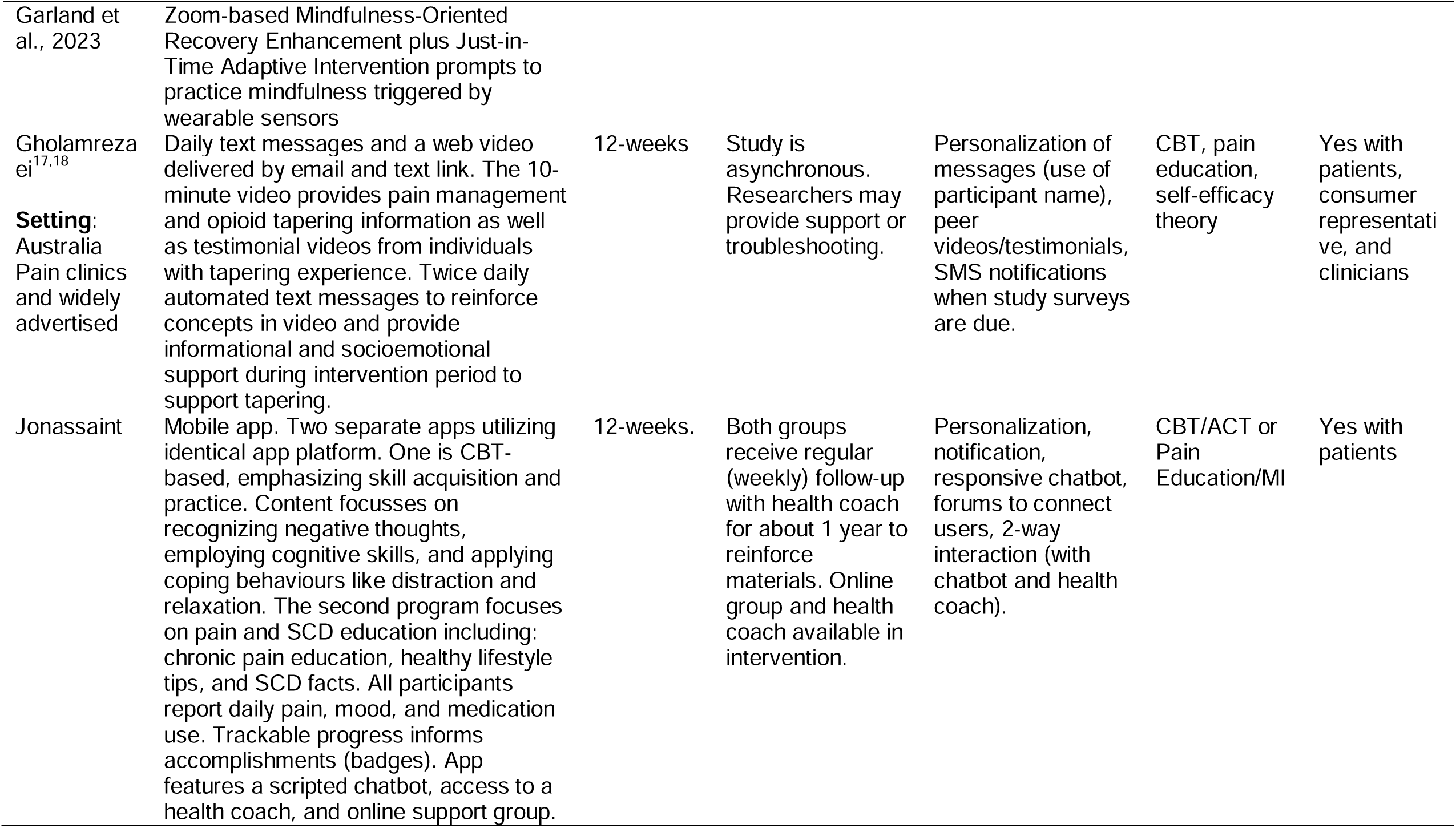
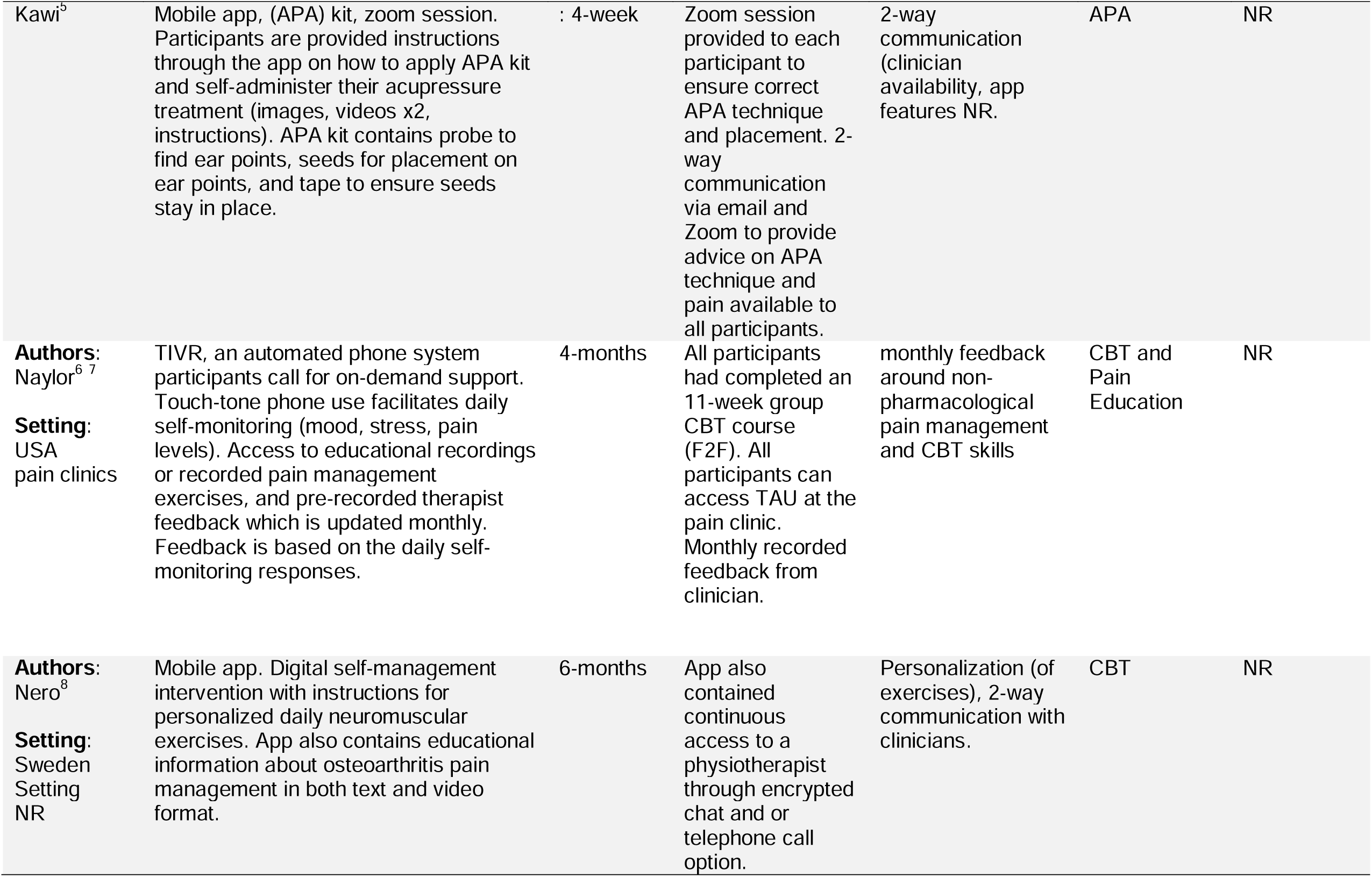
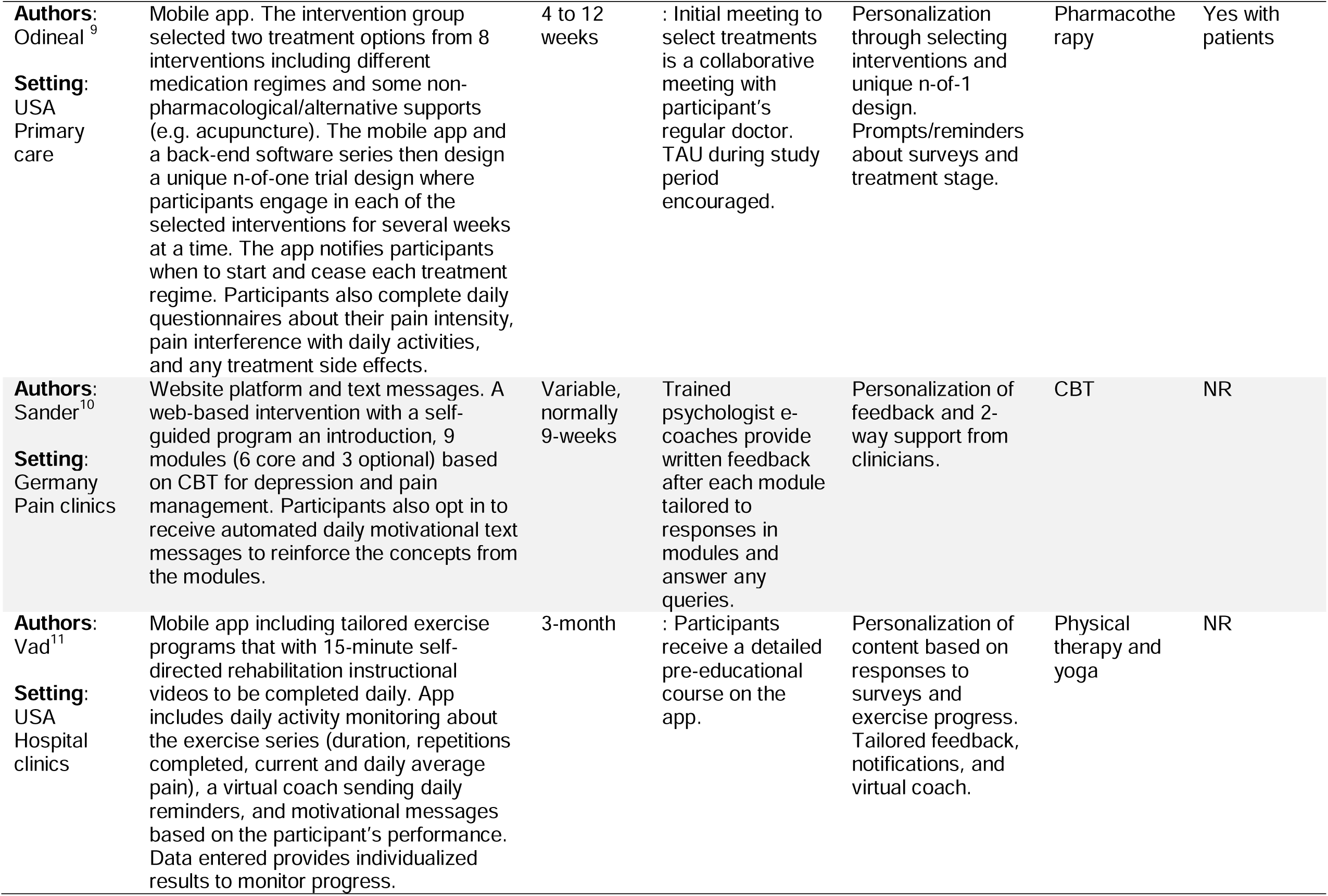
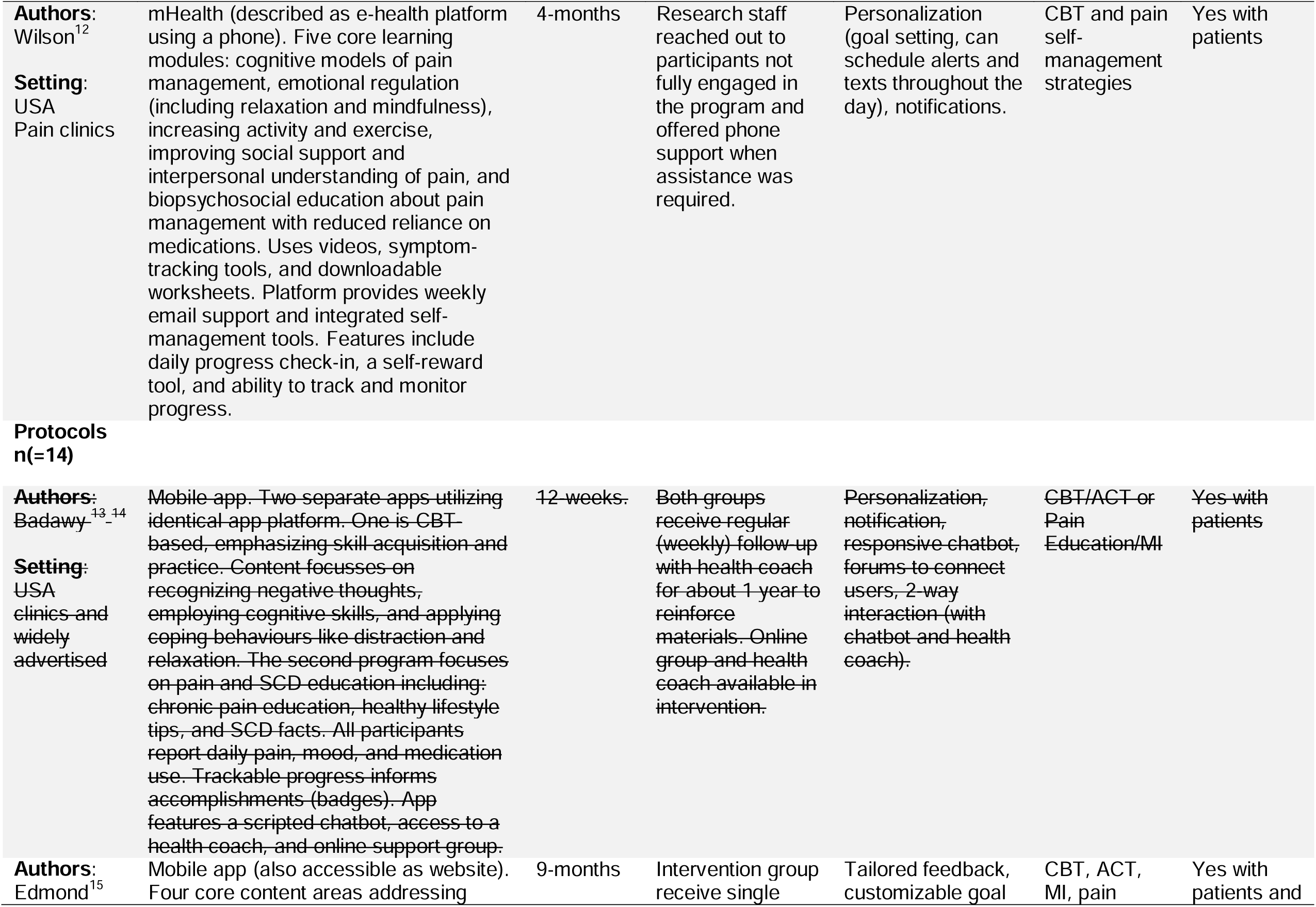
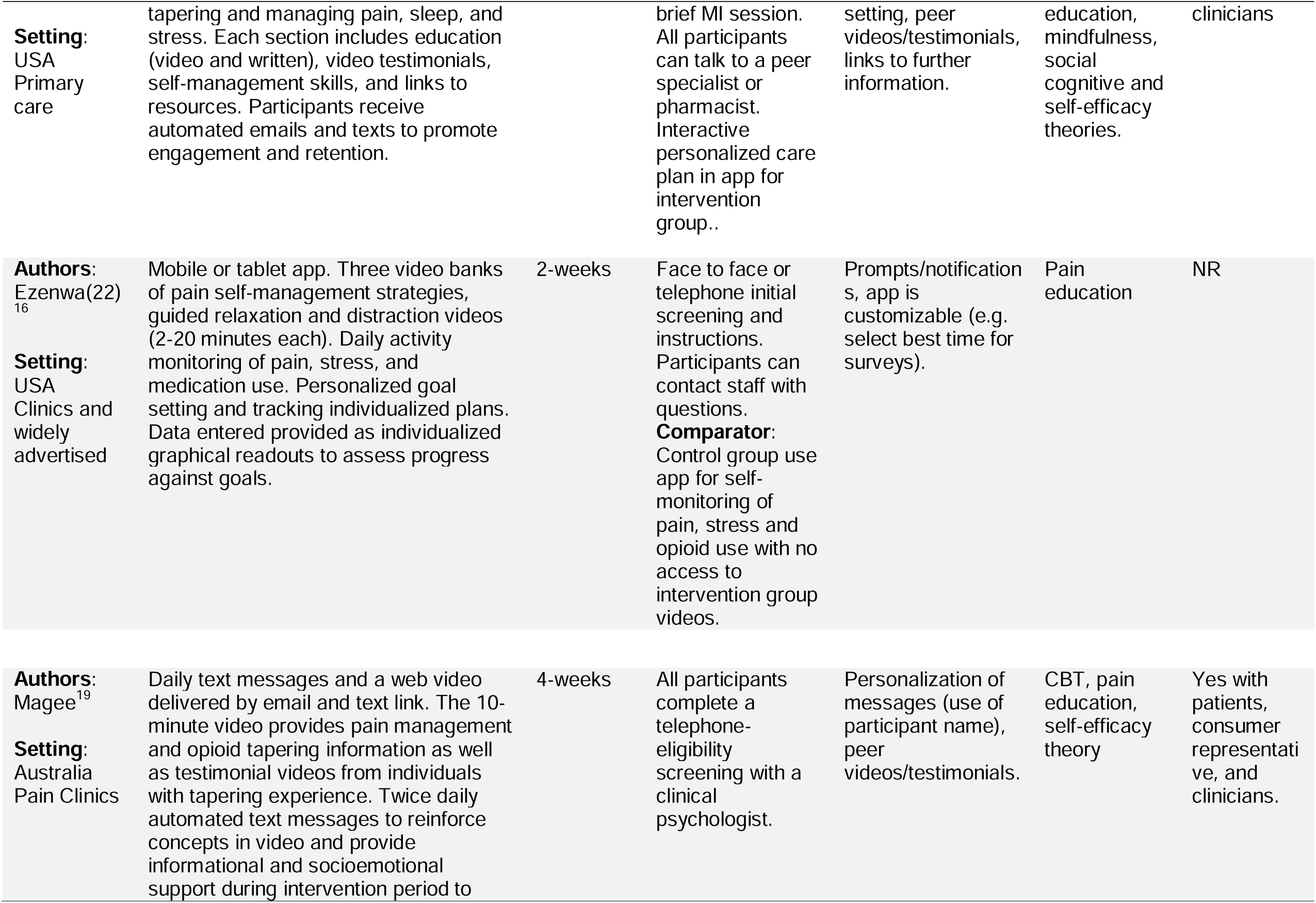
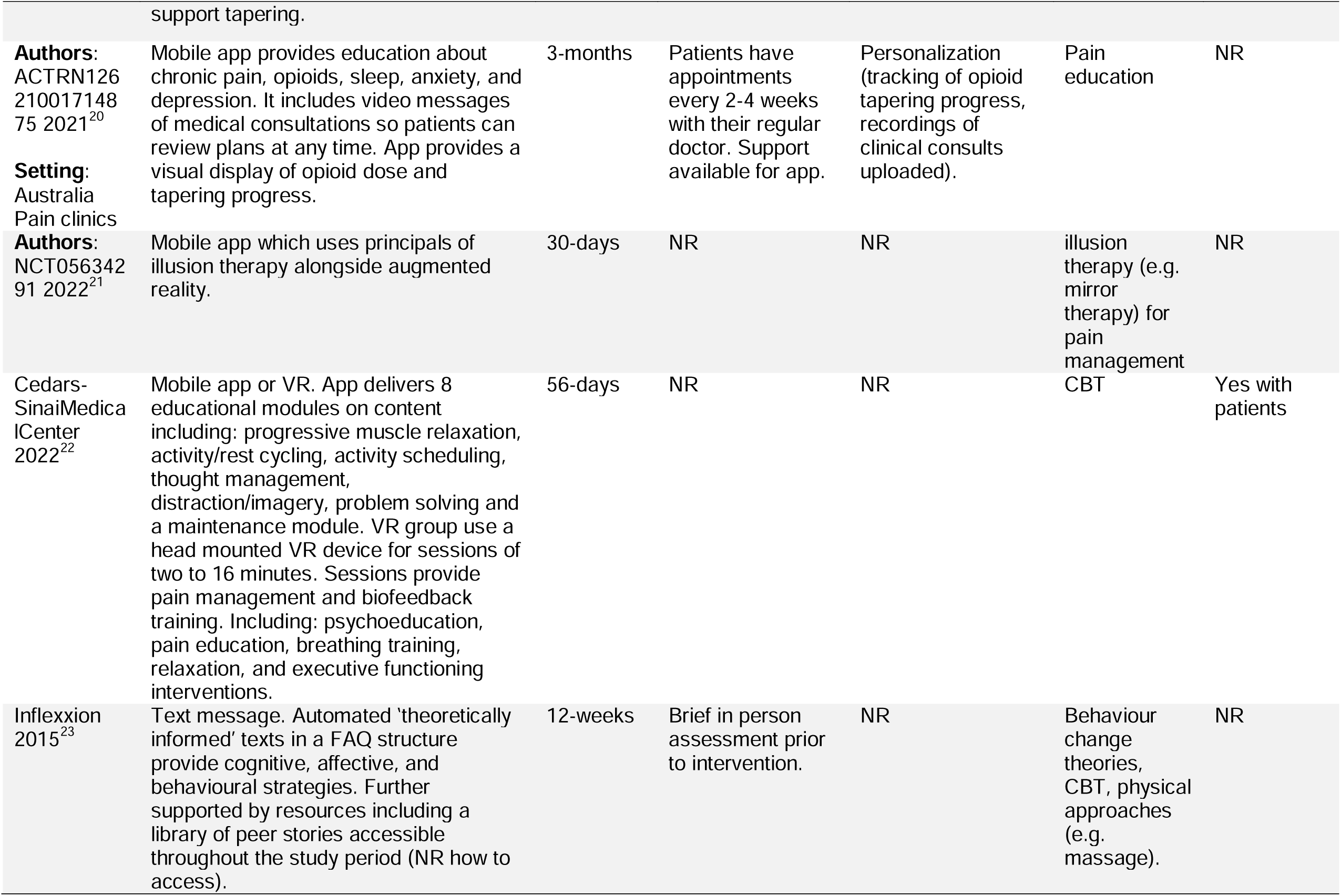
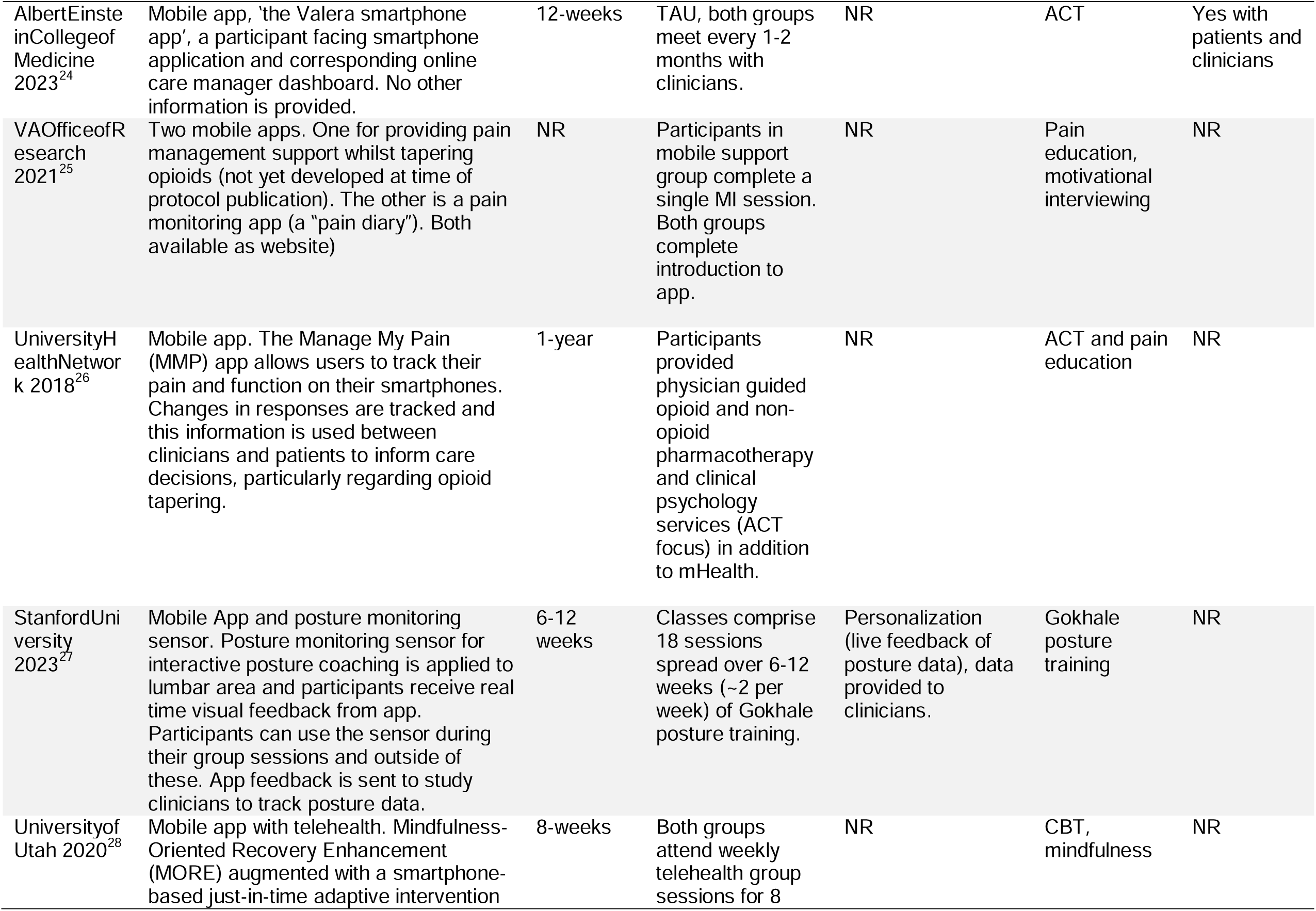
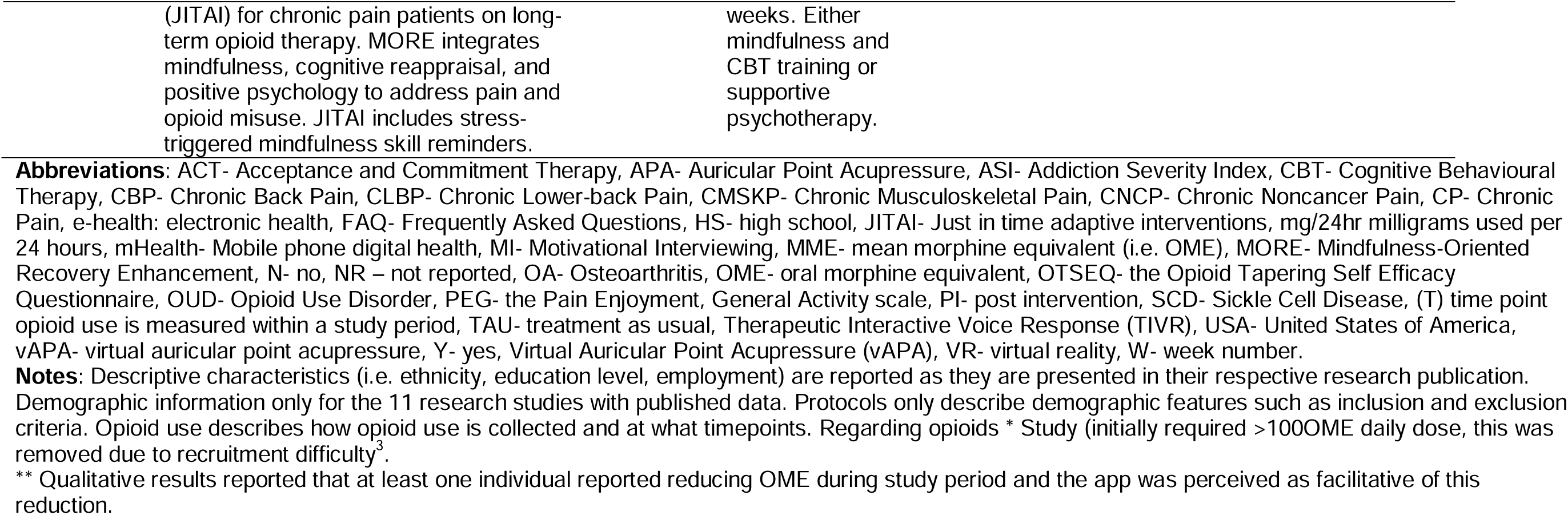
Intervention characteristics in the reviewed studies (n=25 studies)

### In what geographical and clinical contexts (research settings) have the mHealth interventions included in the review been delivered?

The published studies were conducted in the USA^44-47,49,51,52^, Canada^43^, Germany^50^, Australia^42^, and Sweden^48^. The research protocols reported studies to be conducted in the USA^55-58,61-66^, Canada^67^, and Australia.^54 60 59^ The most common recruitment setting was pain clinics, where five research studies^43 42 50 52 47^ and five protocols ^54 64 60 62 67 65^ recruited participants. Three articles, two research studies^45,51^ and one protocol^61^, recruited from other hospital clinics including: sickle cell disease (SCD) clinics, a surgical unit, and a buprenorphine treatment clinic for opioid use disorder (OUD)^61^. Five articles, two research studies^44 49^ and three protocols^63 57,66^, recruited from primary care. Five articles, one research study^46^ and four protocols^59 55,58 55^ reported a hybrid strategy, recruiting from both medical clinics and through wider community advertising. Two articles, one conference abstract ^48^ and one protocol^56^, did not report their recruitment setting.

### What are the characteristics of the study sample populations?

The sample populations were diverse/heterogeneous. Participants of the 25 articles had a range of chronic pain conditions. Twelve of the investigations, six research studies and six protocols, focussed on participants with any chronic non-cancer pain (CNCP) condition including several sub-populations. These subpopulations included patients with difficulty pacing^42^, patients who had recently completed an 11-week group Cognitive Behavioural Therapy (CBT) pain program^47^, participants living in regional areas^47,56^, individuals with sub-diagnostic depression symptoms and CNCP^50^, participants with CNCP and an upcoming surgery^67^, participants with CNCP and comorbid OUD^61^, and participants receiving buprenorphine and opioids^44^. Two studies reported high proportions of participants experiencing comorbid substance use disorder^44,46^. Other pain conditions included, chronic pain from arthritis^48,62^, chronic lower back pain^50,51,65^, chronic pain due to SCD^45 55,58^, and chronic musculoskeletal pain^47^. Four of the protocols did not specify the chronic pain condition^54,57,63,64^. Four studies (all from the USA), one research article^44^ and three protocols^57,63,66^, recruited veterans.

### Demographics

The data extraction table (Table 3) lists demographic variables where reported. Participant ages across all studies had a mean of 49.4 years (SD= 10.7, Range= 26.9 to 63). All studies reported gender as a binary variable. The proportion split of gender or sex was variable, with the percentage of females ranging from 7.5%^44^ to 96%^47^. Eight (73%) of the research studies reported some sample demographic information related to ethnicity, education, and employment status^36 44,45,49,50,52 41 46^. Ethnicity was an inclusion criteria in one SCD study^45^. Education level was reported in eight of the 25 articles^42,44-47,49,50,52^. Relationship status was reported in four studies^42,44,49,50^. Employment data was reported in six studies^42-44,46,49,52^. Only three research protocols specified what demographic or population statistics would be collected^47 60 59^. Whilst no research protocol planned to restrict recruitment based on gender, two of the 14 protocols had ethnicity requirements. One aimed to recruit mostly participants of African descent who live with SCD^58^. The other protocol only planned to recruit Black and Hispanic participants, identified as a marginalised population in the study catchment area^61^.

**Table 3.**
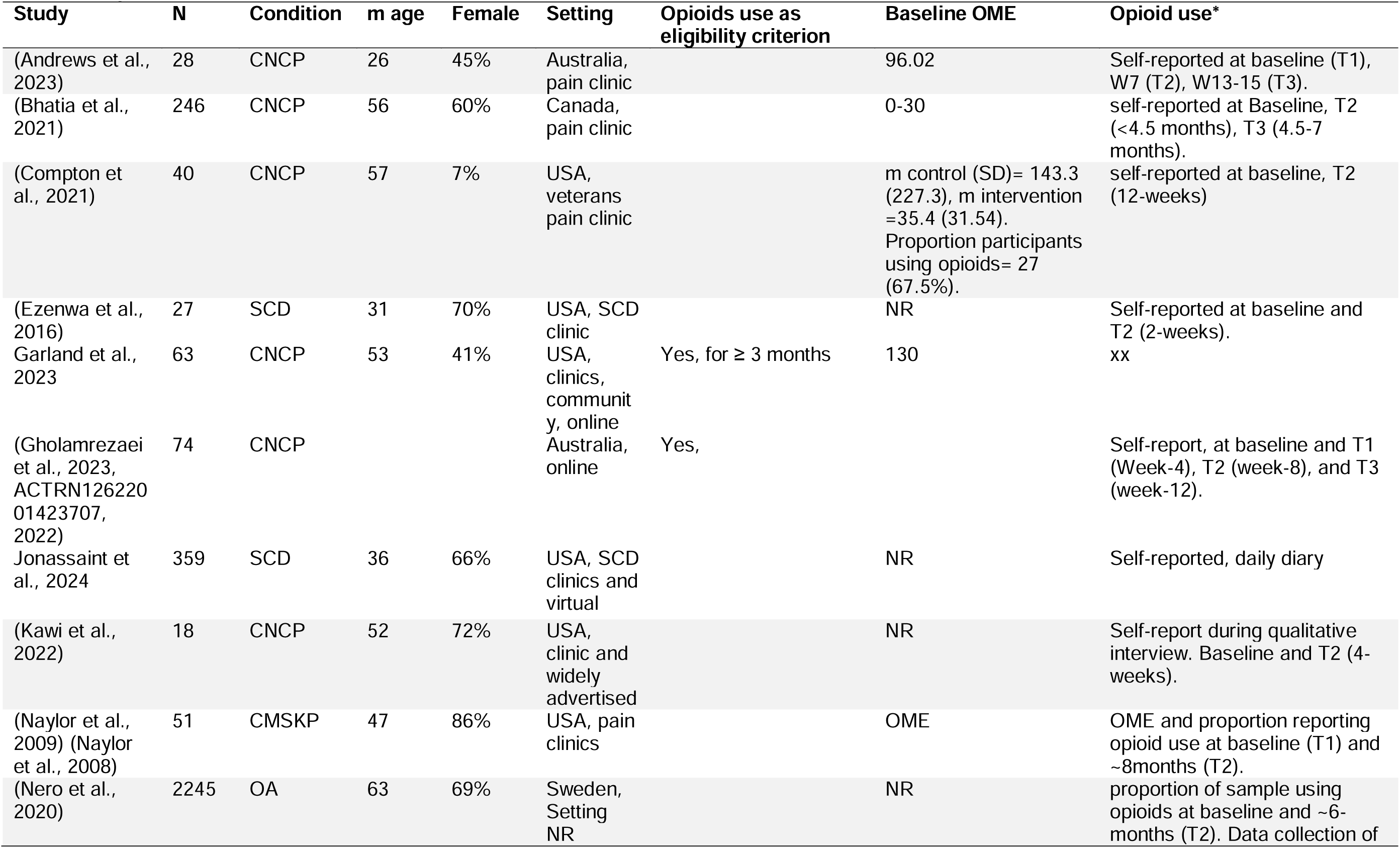
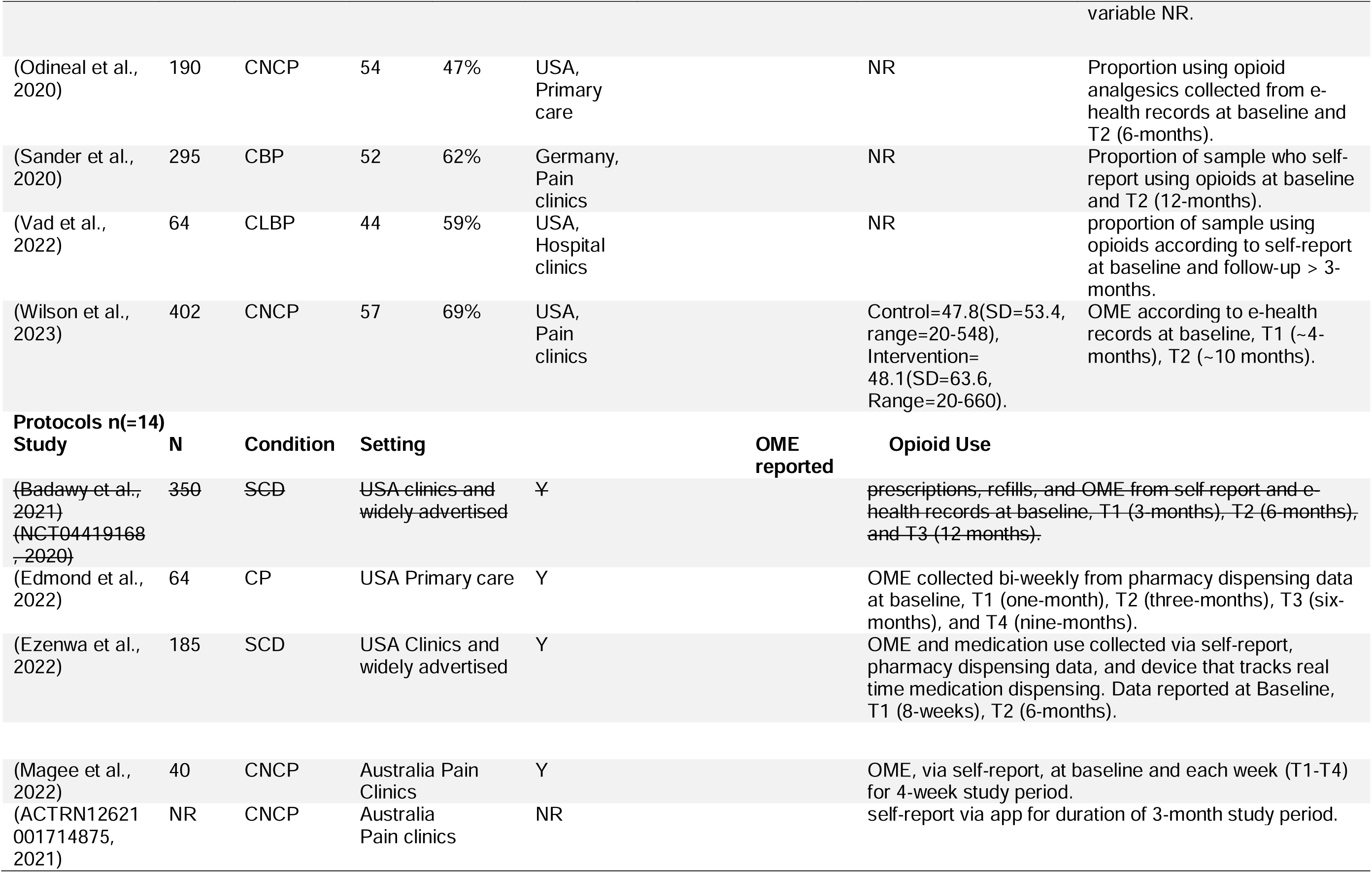
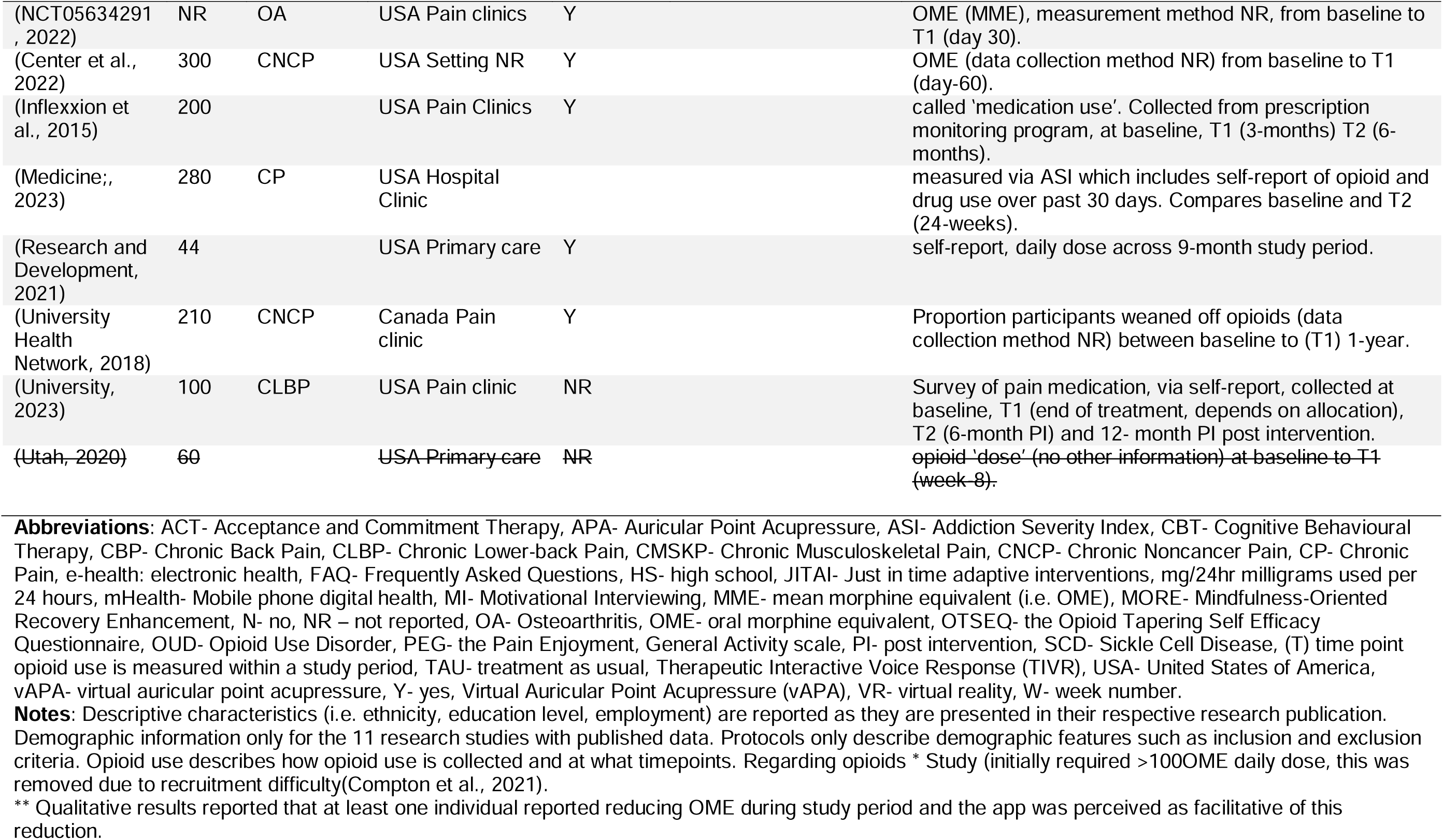
Population characteristics in the reviewed studies.

### Opioids

Taking prescribed opioids was an inclusion criteria in 13 articles, two research studies^44,52^ and eleven protocols^57 54 55 58 60 59 61 62 63 67 66^. Four articles, one research study^45^ and three protocols^54 60 59^, only recruited participants with chronic pain who were tapering opioids. Four articles (two research studies^44,52^, two protocols^60 56^), had specific oral morphine equivalent dose (OME) requirements as part of their eligibility criteria. Of these, two were set at 20mg OME^56 41^. To increase recruitment one study abandoned their dose requirement^44^ and a protocol was amended to reduce daily OME from 60mg to 30mg^60^. Some of the articles reported required minimum durations participants must take opioids, ranging from four weeks^59^ to up to 90 days^55^. The proportion of the sample using opioids was the primary outcome in one^56^ and opioid dose was the primary outcome in seven^49,52,57,61-63,67^ (28% of total) articles that required participants to be using opioids. Three articles that required participants to use opioids did not have opioid dose or use as the key opioid outcome measure. Of these studies the primary outcomes were opioid tapering self-efficacy (two protocols)^60 59^ and abnormalities in eye movement throughout the opioid withdrawal process (one protocol^54^).One article required participants to take opioids, but the primary outcome was physical activity^33^. Of the one published research article requiring participants to take opioids, which reported OME as the primary outcome measure (alongside pain) the authors report 105 or 196 participants (53.6%) receiving their four-month mHealth intervention were able to reduce at least a 15 percent reduction in daily OME compared to 85 of 201 (42.3%) of treatment as usual participants, which the authors report as statistically significant^52^.

Across the literature included in the review the most common ways of reporting opioid use was opioid dose, mostly reported as OME (five studies^42-44,47 46^, 10 protocols^55 57 58 60 59 62 56 64 63 66^), and the proportion of the sample reporting using any opioids (five research articles^45,48-51^, one protocol^67^). Some studies included both OME and sample proportions using opioids (two research articles^44,47^ one protocol^56^). One research study reported opioid outcomes within qualitative interviews only^46^. Self-report was the most common method of data collection, used by six research studies and five protocols^54,60,61,63,65^. Five articles, two research studies^49,52^ and three protocols^55,67 59^ used e-health records. One protocol used pharmacy dispensing to track opioid use, self-report, and a device which tracks real-time medication access^58^. Notably, one research study^48^ and five protocols^54,56,66,67 62^ (25% of results) did not report how they collected opioid data.

Several investigations had additional opioid-related outcome measures including: opioid tapering self-efficacy^60 59^, the Current Opioid Misuse Measure (COMM) to monitor indicators of aberrant drug-related behaviours^55,66 64^; the Screener and Opioid Assessment for Patients with Pain-Revised (SOAPP-R) to evaluate risk for developing problems on long-term opioid use^64^; participant experiences of opioid withdrawal symptoms^60 59^, the Addiction Severity Index (ASI) an assessment of drug use in the past 30 days^61^; opioid craving using the Medication Craving Scale^61^; and momentary craving^66^.

### What are the characteristics of the mHealth interventions in the reviewed studies? Modes of delivery

Mobile applications (apps) were used in 20 of the 25 articles (eight research studies, 12 protocols). Most of these studies only used an app, but some had additional features such as activity monitors^42,44^, supportive text messages^44^, posture monitoring sensors^65^, and an acupressure kit and video conference sessions with an acupressure clinician^46^. One protocol planned to compare the use of a mobile app with a virtual reality (VR) intervention for chronic pain^56^. One research study used Therapeutic Interactive Voice Response (TIVR)^47^, a system which uses touch tone features during a call to provide pre-recorded support. Four articles (one research study, three protocols) predominantly used text messaging for their mHealth intervention. These research studies included a website platform with text message support^50^, twice daily text messages with a 10-minute video at study commencement^60 59^, and one protocol provided text messages only^56^.

### Degree of Clinician Contact

Clinician contact was varied. Five articles (three research studies^42,61,67^, two protocols^65,66^) had clinician delivered components alongside their digital support. These included appointments with a regular clinician^61^, physician and clinical psychology sessions^67^, twice weekly posture training classes for 6-12 weeks^65^, and weekly psychology telehealth sessions for eight weeks^66^. Five articles (three research studies^46,48,50^, two protocols^55,57^) reported clinician support embedded within their mHealth interventions. These included telehealth consultations and email support^46^, written feedback from clinicians delivered into app^50^, digital chat in app with request telephone call feature^48^, online peer groups and online health coach^55^, and on-request clinician or peer support^57^. Eight articles (four research studies^44 45 49,51^, four protocols ^60 63 64 58^) had clinician interaction at the start of the intervention including demonstrations and eligibility assessments^64 60^, and a single session of motivational interviewing^63^. Five articles (two research studies^43,47^, three protocols ^54 52 59^) reported no direct clinician contact. However, most of these studies specified they would provide technical support or follow up if needed. Two protocols^56,62^ did not report the degree of clinician contact.

### Tailoring and engagement strategies

A range of strategies were reported which can be considered tailoring and engagement. Sixteen of the 25 publications (nine research studies ^42 43 44,47 48 49 50 51,52^, seven protocols^55 57 58 60 59 54,65^) reported some degree of personalisation within their digital health interventions. For example, two studies specified use of participants’ names in text messages to increase engagement^60 59^. More sophisticated personalisation included using interactive charts and graphs to visually present data that participants input into their mHealth platform^42 43,54,65^. A range of articles described providing personalised feedback through their apps^50,51,57^ or through pre-recorded voice messages from clinicians^47^. Some interventions allowed participants to customise times of days they would complete surveys or receive alerts^52,58^. An exercise-focused intervention provided personalised exercise programs^48^. Reminders, prompts, and push notifications were specifically mentioned as strategies to increase adherence and engagement in nine articles (six research studies^42,43,45,49,51,52^, three protocols^55 47 59^. Six articles (five research studies, one protocol) specified the use of two-way communication within their interventions. This included clinicians being able to see data from the app^43,65^ and live clinician communication (either through the app or a request contact function)^46,48,51^. One protocol described multiple communication methods including a responsive chatbot and forums to connect users^55^. Three research articles included the use of rewards, one using a variable interval reward schedule with financial rewards^44^. The other two studies provided a device for the study period, which could be a potential incentive^42,45^. Three protocols reported providing peer testimonials in the form of videos^60 59 57^. Seven protocols^62 56 64 61 63 67 66^ did not report any intervention components which were considered engagement strategies.

### What is the degree to which consumer involvement or co-design is included in the content development, intervention, and study design of the reviewed studies?

There was a range in detail describing co-design. Fourteen of the 25 articles (six research studies^44 50,51 46 48 47^ and eight protocols^58 54 62 64 63 67 65,66^) did not report or specify any information on co-design. Eleven articles (five research studies^45 49 43 42 52^ and six protocols^57 60 59 55 56,61^) did specify some degree of co-design. Of these articles, ten (four research studies^45 49 42 52^ and six protocols ^60 59 57 55 56,61^) described co-design with patients. This included soliciting feedback from patients about the intervention in design phases^45^. Seven of the articles (three research studies^43 42^ and four protocols^57 60 59 51^) described co-design with clinicians, such as surveying clinicians about the perceived acceptability and appropriateness of intervention content^60 59^. In another study, clinician co-design was used to integrate the digital health platform into clinical workflows within a hospital^43^. Three articles reported co-design with other stakeholders. One study considered their software engineer as a co-design member^42^. Two protocols reported co-designing their intervention with a representative of a consumer advocacy organisation^60 59^. Some protocols reported plans to follow up their studies with separate investigations of user/participant experiences through qualitative interviews with participants^43,46,51,67 60 59^. Five articles (two research studies^42,52^, three protocols^62 56,64^) reported affiliations with a government service. Eight studies (two research studies^43,51^, four protocols^55 56 54,66^) reported affiliation with an enterprise or business. These mostly included staff of the companies which owned the digital health interventions used in their study^43,54,66^. Four of the protocols reported affiliations with consumer representative organisations^55 64 60 59^.

### What is the degree to which theoretical frameworks and clinical interventions underpin the design and or content of mHealth interventions?

Only one study, a conference abstract^48^, did not specify the theories, frameworks, or clinical interventions underpinning their mHealth intervention. One research article intervention primarily focussed on pharmacotherapy^49^. Most of the articles reported their intervention content drew upon at least one psychological approach including: CBT (eight total, three published studies^50 47,52^, five protocols ^60 59 55 56,66^), acceptance and commitment therapy (ACT; four total, one published study^43^, three protocols^55 61,67^), mindfulness training (one protocol^66^), behavioural incentives theory (one research study^44^), motivational interviewing (three protocols^57 55,63^), social cognitive theory and self-efficacy theory (three procotols^57 60 59^), and pain self-management education (11 total, four published studies^45 42 47,52^, seven protocols^58 60 59 55 63 67 54,62^), which was the most common. Physical approaches were also referenced including: yoga and pilates (one research study^51^), physiotherapy and exercise (one research study^52^), and Gokhale posture therapy (one protocol^65^). Other physical approaches included massage, chiropractic and acupuncture approaches (one procotol^64^) and auricular point acupressure (APA; one research study^46^).

### What is the scope of evidence on acceptability and feasibility of interventions in the reviewed studies?

Of the 25 included articles, eight (four research studies^42 45 46,51^, four protocols^58 57,63 60^) reported acceptability or feasibility as the primary outcome of the study. Another eight (two research articles^47,48^, six protocols^54 62 61 63 66,67^) reported neither acceptability nor feasibility outcomes in their reported or planned data. Seventeen articles (nine research studies^42 44 45 49 43 51 46,50,52^, eight protocols^58 57 55 64 56,65 60 59^) reported acceptability outcomes. The variables used to evaluate acceptability included adherence/use (four research studies^43 42 51,52^) one article reported 111 of 176 (63.4%) participants used the intervention for at least 30 days, indicating app acceptability ^43^. Another study reported 52% of participants were compliant (using the app at least three times per week over 12-weeks), while 65% of the 75 participants rated their app experience as good or excellent^51^.Another adherence measure used a 0-6 scale with scores of two or more correlated to sufficient completion of educational content proven to provide benefits in previous research^52^. Of 200 participants, 136 (68%) of participants scored at least two (m=2.5, SD=2.0)^42^. Questionnaire responses (two research studies^45,50^, three protocols^58 46 60^) included measures such as a six-item study acceptability scale, 13 item tablet (device) acceptability scale, and a six-item open ended questionnaire which was completed as an interview^45^. The results showed participants were satisfied using portable digital devices for data collection and accessing mHealth interventions^45^. Another study utilised the Client Satisfaction with online Interventions Questionnaire-8 (8-item, range 8-32)^69^ where participants scored a mean of 22.87 (SD=4.92)^50^. End of-study interviews (three research studies^42 45,46^, eight protocols^58 57 55 64 56,65 60 59^) generally reported reasonable or good acceptability^42^. One study used semi-structured interviews to establish acceptability and provided many quotes^46^. Dropout rates (two articles^44,49^), indicated 95% (38 of 40) completion in one ^44^, and 6% dropout rates (14 of 215) in another, with 5% (11 of 215) also not authorising follow-up access to medical records^49^.

Feasibility, was included as an outcome in 14 studies (six research articles^42 45 50 51 46 52^, eight protocols^60 59 57 58 55 65 57,61^) and not reported in four research studies^44 49 47 48^. Feasibility measures included enrolment success(protocol^58^), dropouts (protocol^60^), missing data (protocol^60^), cost effectiveness (protocol^61^), and intervention use during (two procotols^55,58^) and after the study period (three protocols^58 57,65^). One study assessed feasibility by clinic attendance, with 63% of participants rescheduling appointments, and four of 16 participants repeating the one-week monitoring period with an activity tracker^42^. Another study reported a 96% completion rate of baseline assessments, with all 27 participants completing 100% of the two week data collection despite recruitment challenges including some participants being to ill to complete their data collection^45^. Another study used interviews to evaluate feasibility^46^. Adverse events were the most common feasibility variable, in nine articles (six research studies^50 51 42 45,46 52^, three protocols^59 55,65^). Most studies reported no adverse events^51 42 43 46 44 49 47 48^ and collected adverse event data retrospectively, though some monitored for adverse events during study period^42 50^. In one study a significant increase in pain or psychological distress was considered an adverse event, occurring for 42.5% of mHealth and 47.0% of control participants, though no serious adverse events were reported^52^. One study terminated a face to face assessment (and the participants study participation) due to a patient becoming agitated and unresponsive to hospital staff^45^. In the largest study (n=9757), 1.3% (129 participants) reported adverse events^50^, with no differences between intervention and control groups, no adverse events attributed to the intervention, and no serious adverse events such as hospitalisation or suicide attempts^50^.

## Discussion

To identify the scope of research on mobile phone digital health (mHealth) interventions to support opioid tapering, this study identified and described 25 research publications (research studies and protocols) recruiting adults with chronic pain which report changes in opioid use, before and after an mHealth intervention. Regarding the geographical context, participants were from five countries, with approximately half (13 of 25) conducted in the USA. Most articles recruited participants from health services (e.g. pain clinics and primary care). Most studies were published during or after 2021, potentially reflecting the increase in digital health interventions in recent years in response to technological advancement and health servicing needs during COVID-19^70^ and the increased need for tapering support in chronic pain^13^. All participants had chronic pain and nearly half the research recruited participants with any chronic pain condition. Other demographic features were variable across studies. Some subgroups emerged including veterans (3 articles) and adults living with sickle cell disease (3 articles). Notably, where reported, the proportion of samples not working was between 40 to 60 percent.

### Opioids

Most of the articles, required participants to use ongoing prescription opioid medications. However, only two of these articles were published research studies (i.e. not protocols). Of the other studies that reported data, only a subset of each sample took opioids, potentially limiting the conclusiveness of any results. Participant self-reported medication use was the most common form of collecting opioid data (typically reported as OME). Some articles used e-health records and pharmacy dispensing data to monitor medication use. Self-report is prone to error but is a feasible long-term method of data collection^59^ particularly for tapering, often a long process^8,71^. The second most frequent variable after OME was the proportion of samples self-reporting using any opioids (binary variable). The proportion of a sample reporting opioid use is a useful statistic, particularly for samples where not all participants take opioids, or to evaluate tapering success in larger population interventions^13^. However, OME is more clinically relevant. Opioid risks are dose specific so reduction without cessation is a meaningful goal^13^ and enables contrasting how dose may interact with sleep, pain and mood scores^13,15^, data regularly collected within many mHealth interventions in this review. Several included articles used evidence based measures for opioid related outcomes including drug-related behaviours^55,66 64^; risk for developing problems on long-term opioids^64^; withdrawal symptoms^60 59^, drug use^61^; and opioid craving^61,66^. Finally, two interventions used opioid tapering self-efficacy as their primary opioid related outcome, hypothesising improved self-efficacy would facilitate tapering^60 59^.

### mHealth

Apps were used in 20 of the 25 articles and there were five included articles which used text messaging (one of which was an app with optional texts). One study used interactive voice recordings within a call. There was high variability in the characteristics of the mHealth interventions. The design and features of the mHealth interventions were linked to the goals of each study. All interventions had some fully asynchronous mHealth support, but many interventions included group or individual meetings and online access to peers or clinicians. Resources required were variable and some required external devices including VR equipment and activity trackers. The mHealth interventions often embedded features to improve engagement including personalisation and interactive communication.

Underpinning most mHealth interventions were theoretical frameworks and clinical interventions. Mostly these were psychological approaches, including CBT and pain education, or physical activity programs accompanying a psychological approach. Despite interventions being designed for specific subgroups this conceptual overlap may reflect that the pain management strategies most suitable for non-pharmacological approaches (i.e. they support tapering and can be delivered digitally) are pain self-management and psychologically oriented interventions^72 73^. However, none of the published studies reported that they tried to evaluate the hypothesised mediating mechanisms (e.g. self-efficacy and correlations with tapering outcomes).

Further, consumer involvement is likely to enhance: consumer perceptions towards interventions (e.g. relevance), successful recruitment, and acceptability and feasibility of clinical trials^70 74 75,76^. In our research, the only articles specifically investigating tapering support with text message interventions^59,60^, extensive co-design research was conducted to suit the specific consumers^15 77^. Despite these benefits fourteen articles did not report any co-design information and many studies described co-design in a limited way.

### Limitations

This review only included chronic pain studies and excluded similar mHealth studies for cancer, acute, and post-surgical pain. Further, the mHealth interventions must be deliverable via mobile phones. Therefore, we excluded studies that used computer based or website-based interventions. Digital interventions which may have worked on a smart phone but were described as computer-based may have been excluded. Despite these limits, we considered our design was the most appropriate and feasible for the focus of this investigation, the scope of mobile phone and chronic pain research with tapering outcomes.This scoping review did not evaluate the methodological quality of the studies or the quality of the data^34^. The methodological lack of data appraisal in scoping reviews should be considered in interpreting the results. The diversity in mHealth interventions, outcome measures, and reporting also necessitates caution when interpreting the overall results.

### Future directions

Regarding the articles that were included in the review, eight were pilot studies. Not all participants in the published research took opioids, and those studies range in opioid dose, demographic characteristics, and the degree of care they received. With 14 protocols included in this study more data is in the pipeline. But may prove not to be completed, which may change the scope of these findings. At this stage the limited range of quantitative opioid data described in the results prevents accurately comparing the effectiveness of each interventions opioid data. Meaning systematic review of results is not yet necessary. But in time, may enable comparison of outcomes across the study populations and intervention types, which may help evaluate effectiveness.

Dropouts occurred in most completed research, with some studies using tracking and follow-up methods for safety concerns, such as increased pain and psychological distress, treated as adverse events requiring clinician follow-up^50 52^. This demonstrates strong support in some studies. But such assertive care may not be feasible in larger studies or scalable mHealth programs, limiting mHealth’s ability to address clinician shortages and offer cost-effective, accessible support^22-24^. Most studies concluded their interventions were generally safe and acceptable. However, if distress and pain, expected in chronic pain and opioid tapering, should be classified as adverse events is debatable^32^. Future research should consider safety, monitoring, and outcomes for participants undergoing tapering whilst receiving digital support^32^.

Researchers seeking to develop scalable mHealth supports for tapering amongst chronic pain populations could inform their future work with some proposals for consistency. Standardised opioid doses should be used to report opioid use^13^. Where it is not a study requirement to take opioids, the proportion of a sample using opioids can be reported in addition to OME^13^. Given the reported recruitment difficulties^44,60^ of specific OME requirements, feasibility may improve if studies include any participant taking opioids for chronic pain regardless of their initial dose^13^.

The mHealth interventions and their features were varied. Demonstrating the flexibility that digital health and mHealth supports can provide. Whilst there was some conceptual overlap, the degree of co-design, design features to improve engagement, and the level of clinician support are some of the variable intervention features that could be evaluated for their impact on pain and tapering outcomes. Particularly where these features could limit an mHealth support being acceptable, scalable, or affordable in another setting. Larger RCTs and implementation studies will provide more data regarding resource demands, implementation challenges, and to evaluate superiority of any approach^70^.

### Summary and Conclusions

The rapid growth of digital tools for managing chronic pain and related opioid use has outpaced the availability of evidence-based interventions^78^. This scoping review examined the research investigating mHealth interventions supporting opioid tapering in chronic pain management, identifying 25 studies, including 14 protocols and 11 completed studies. Most interventions were app-based, though text messaging was also common. Psychological frameworks such as CBT, ACT, and self-efficacy theory underpinned most of the mHealth interventions. Early findings suggest that mHealth interventions are generally safe, with low rates of adverse events, and acceptable to those with chronic pain who may be reducing or seeking to reduce their opioid use. However, the limited scope of the completed studies, along with a wide range of mHealth designs, outcome measures, and few published articles specifically focussing on opioid tapering prevents clear conclusions about the most effective, if any, mHealth tools for opioid tapering. As more data from the 14 protocols and any other studies becomes available, it will be possible to better evaluate the feasibility, acceptability and efficacy of the mHealth interventions, particularly regarding their impact on supporting individuals living with chronic pain to reduce their reliance on opioid medications. These results demonstrate the breadth of options that are available to researchers, clinicians, and consumers to support tapering.

## Supporting information

Appendix - Search Strategy

Appendix PRISMA-ScR

## Data Availability

All data produced in the present study are available upon reasonable request to the authors

## Contributors

MM wrote the first draft of this manuscript and protocol, led the search and extraction. AG, AM, and AS supported the search and screening. MM, CAJ, AG finalised the data extraction. All authors contributed to the study conceptualisation, development of the manuscript, and agreed to the final version of this manuscript to be published.

