## Appendix - Search Strategy for "Reducing opioid use for chronic pain with mobile health interventions: a systematic scoping review"

| **APA Psych Info Search (Via OVID) Conducted 23/4/23** | |
| --- | --- |
| **Results** | **Type** |
| 1 | Chronic Pain.mp. or exp Chronic Pain/ |
| 2 | exp Musculoskeletal Disorders/ or exp Pain Management/ or exp Chronic Pain/ or Musculoskeletal Pain.mp. |
| 3 | Pain Management.mp. or exp Pain Management/ |
| 4 | (("Pain" or "pain*") and ("chronic" or "long term" or "long-term" or "ongoing" or "persistent")).mp. [mp=title, abstract, heading word, table of contents, key concepts, original title, tests & measures, mesh word] |
| 5 | 1 or 2 or 3 or 4 |
| 6 | Cell Phone.mp. or exp Mobile Phones/ |
| 7 | exp Smartphones/ or smartphone.mp. |
| 8 | exp Telephone Systems/ or exp Smartphones/ or exp Mobile Phones/ or exp Telemedicine/ or exp Mobile Applications/ or phone*.mp. |
| 9 | exp Telephone Systems/ or telephone*.mp. |
| 10 | exp Mobile Phones/ or cellphone*.mp. |
| 11 | exp Smartphones/ or exp "Smartphone Use"/ or exp Mobile Phones/ or smartphone*.mp. or exp Mobile Applications/ |
| 12 | exp Telemedicine/ or telehealth*.mp. |
| 13 | exp Mobile Applications/ or exp Mobile Phones/ or exp Telemedicine/ or exp Mobile Health/ or mobile healthcare.mp. or exp Mobile Devices/ |
| 14 | exp Mobile Health/ or exp Telemedicine/ or m-health.mp. |
| 15 | mHealth.mp. or exp Mobile Health/ |
| 16 | exp Mobile Health/ or mobile health.mp. |
| 17 | exp Text Messaging/ or exp Messages/ or exp Mobile Phones/ or text message.mp. |
| 18 | exp Text Messaging/ or text messaging.mp. |
| 19 | exp Text Messaging/ or exp Messages/ or exp Mobile Phones/ or SMS.mp. |
| 20 | exp Mobile Phones/ or exp Messages/ or exp Telemedicine/ or exp Text Messaging/ or exp Mobile Health/ or short messaging service*.mp. |
| 21 | short message service*.mp. |
| 22 | exp Mobile Devices/ or exp Mobile Applications/ or Mobile Applications.mp. |
| 23 | exp Mobile Devices/ or exp Mobile Applications/ or exp Mobile Phones/ or mobile application*.mp. |
| 24 | exp Mobile Phones/ or exp Mobile Applications/ or mobile app*.mp. |
| 25 | exp Mobile Devices/ or exp Mobile Applications/ or exp Mobile Health/ or exp Mobile Phones/ or exp Smartphones/ or phone app*.mp. |
| 26 | smartphone app*.mp. |
| 27 | exp Mobile Applications/ or digital app*.mp. |
| 28 | exp Mobile Phones/ or exp Smartphones/ or exp Mobile Applications/ or exp Mobile Devices/ or android app*.mp. or exp Mobile Health/ |
| 29 | iphone app*.mp. |
| 30 | iphone.mp. |
| 31 | exp Mobile Phones/ or exp Mobile Applications/ or android app.mp. |
| 32 | 6 or 7 or 8 or 9 or 10 or 11 or 12 or 13 or 14 or 15 or 16 or 17 or 18 or 19 or 20 or 21 or 22 or 23 or 24 or 25 or 26 or 27 or 28 or 29 or 30 or 31 |
| 33 | opioid.mp. or exp Opioid Analgesics/ |
| 34 | opioid*.mp. or exp Opioid Epidemic/ or exp "Opioid Use Disorder"/ |
| 35 | exp Opiates/ or exp "Opioid Use Disorder"/ or Opioid-Related.mp. |
| 36 | exp Opiates/ or Opiate Substitution Treatment.mp. |
| 37 | exp Methadone Maintenance/ or exp Methadone/ or Methadone.mp. |
| 38 | Buprenorphine.mp. or exp Buprenorphine/ |
| 39 | Fentanyl.mp. or exp Fentanyl/ |
| 40 | Oxycodone.mp. or exp Oxycodone/ |
| 41 | tramadol.mp. or exp Tramadol/ |
| 42 | Codeine.mp. or exp Codeine/ |
| 43 | exp Morphine/ or Morphine.mp. |
| 44 | exp Opiates/ or exp Morphine/ or Hydrocodone.mp. |
| 45 | exp Opiates/ or Hydromorphone.mp. |
| 46 | exp Opiates/ or exp Endogenous Opiates/ or Opiate*.mp. |
| 47 | oxycontin.mp. or exp Oxycodone/ |
| 48 | exp "Opioid Use Disorder"/ or exp Opiates/ or Vicodin.mp. |
| 49 | exp Opiates/ or exp Oxycodone/ or Tapentadol.mp. or exp Tramadol/ |
| 50 | exp Opiates/ or exp Meperidine/ or Pethidine.mp. or exp Morphine/ |
| 51 | exp Meperidine/ or meperidine.mp. |
| 52 | 33 or 34 or 35 or 36 or 37 or 38 or 39 or 40 or 41 or 42 or 43 or 44 or 45 or 46 or 47 or 48 or 49 or 50 or 51 |
| 53 | 5 and 32 and 52 |
